# Adenomas Are Frequent in PMS2 Lynch Syndrome patients but Rarely Mismatch Repair Deficient

**DOI:** 10.1101/2025.09.01.25334843

**Authors:** K.D. Andini, L. Lanjouw, M. Suerink, D. Terlouw, A. Ahadova, S. Aretz, M. Bigirwamungu-Bargeman, H. Bläker, W.H. de Vos Tot Nederveen Cappel, V. Endris, E. Holinski-Feder, M.A.J.M. Jacobs, J.J. Koornstra, A.M.J. Langers, M. Loeffler, A. Lepistö, J.-P. Mecklin, M. Kloor, G. Möslein, P. Peltomäki, K. Pylvänäinen, L. Renkonen-Sinisalo, T.T. Seppälä, C.P. Strassburg, V. Steinke-Lange, M. Morak, R. Hüneburg, S. Redler, C.M.J. Tops, P.C. van de Meeberg, M. van Kouwen, D.B. Vangala, B. Katerberg, M.-L. Verhulst, M. von Knebel Doeberitz, S. Zachariae, C. Engel, H.F.A. Vasen, T. van Wezel, H. Morreau, K. Kok, R.H. Sijmons, M. van Leerdam, M. Nielsen, G. Kats-Ugurlu, S.W. Bajwa-ten Broeke

## Abstract

**Background:** Lynch syndrome (LS) predisposes carriers to the accelerated development of colorectal cancer (CRC). Pathogenic *PMS2* variant (PV) carriers are believed to have the lowest CRC risk and develop MMR deficiency (dMMR) at a later stage of CRC progression. In addition, the *PMS2* adenoma incidence rate is considered lowest among all MMR PV carriers. However, neither the exact adenoma incidence nor the prevalence of dMMR among *PMS2* PV carriers is known.

**Method:** We established a cohort of 171 confirmed *PMS2* PV carriers in the Netherlands and collected clinical excerpts to establish adenoma incidence. We also collected 123 paraffin blocks from *PMS2* PV carriers and investigated PMS2 and MLH1 expression with immunohistochemistry (IHC).

**Results:** We collected 123 paraffin blocks containing adenomatous lesions from *PMS2* PV carriers removed who underwent surveillance colonoscopy between 2018-2023. Of the 123 IHC-stained lesions 109 were tubular adenomas, 86.2% of which (n=94/109) retained PMS2 protein expression. All specimens showed intact MLH1 staining.

**Conclusion:** Adenomas are frequent in *PMS2* PV carriers, although the majority of PMS2-associated adenomas retained MMR-proficiency. The result of our study corroborates the late involvement of *PMS2* deficiency in the evolution of to PMS2 associated CRC.

## Research Letter

Lynch syndrome (LS) predisposes carriers of a germline heterozygous pathogenic variant (PV) in one of the mismatch repair (MMR) genes, *MLH1, MSH2, MSH6* or *PMS2*, to the development of mainly colorectal and endometrial carcinomas^1^. Previous studies reported a relatively low risk of colorectal cancer (CRC) among *PMS2* PV carriers. This has resulted in an underreporting of *PMS2* PV carriers in Lynch syndrome cohorts reported in literature due to clinical recognition and ascertainment, as evidenced by a much higher prevalence of 1:714 of PMS2 variants in the general population compared to other MMR genes^2,3,4^. This suggests that many PMS2-LS families remain undetected by clinical selection criteria. The variation in cancer risk associated with the different subtypes of Lynch syndrome is likely due to differences in molecular pathogenesis of *PMS2*-CRC. Recent reports based on IHC and molecular testing have highlighted three pathways of LS colorectal carcinogenesis^5^. Carriers of *MLH1, MSH2*, and in part *MSH6* PVs may develop LS-associated CRC through MMR-deficient early lesions, such as MMR-deficient crypt foci or MMR-deficient adenomas. In contrast, we have previously reported that *PMS2*-CRC likely develops through MMR-proficient adenoma that transforms into MMR-deficient CRC over time^6^ (Figure 1A). However, our knowledge on *PMS2* carcinogenesis is still limited, since data regarding incidence and characteristics of early *PMS2-*associated adenomas are sparse. Our current study addresses the cumulative incidence and immunohistochemical features of adenomas in *PMS2* PV carriers based on surveillance colonoscopy data on confirmed germline *PMS2* PV carriers registered in the Netherlands.

**Figure 1.**
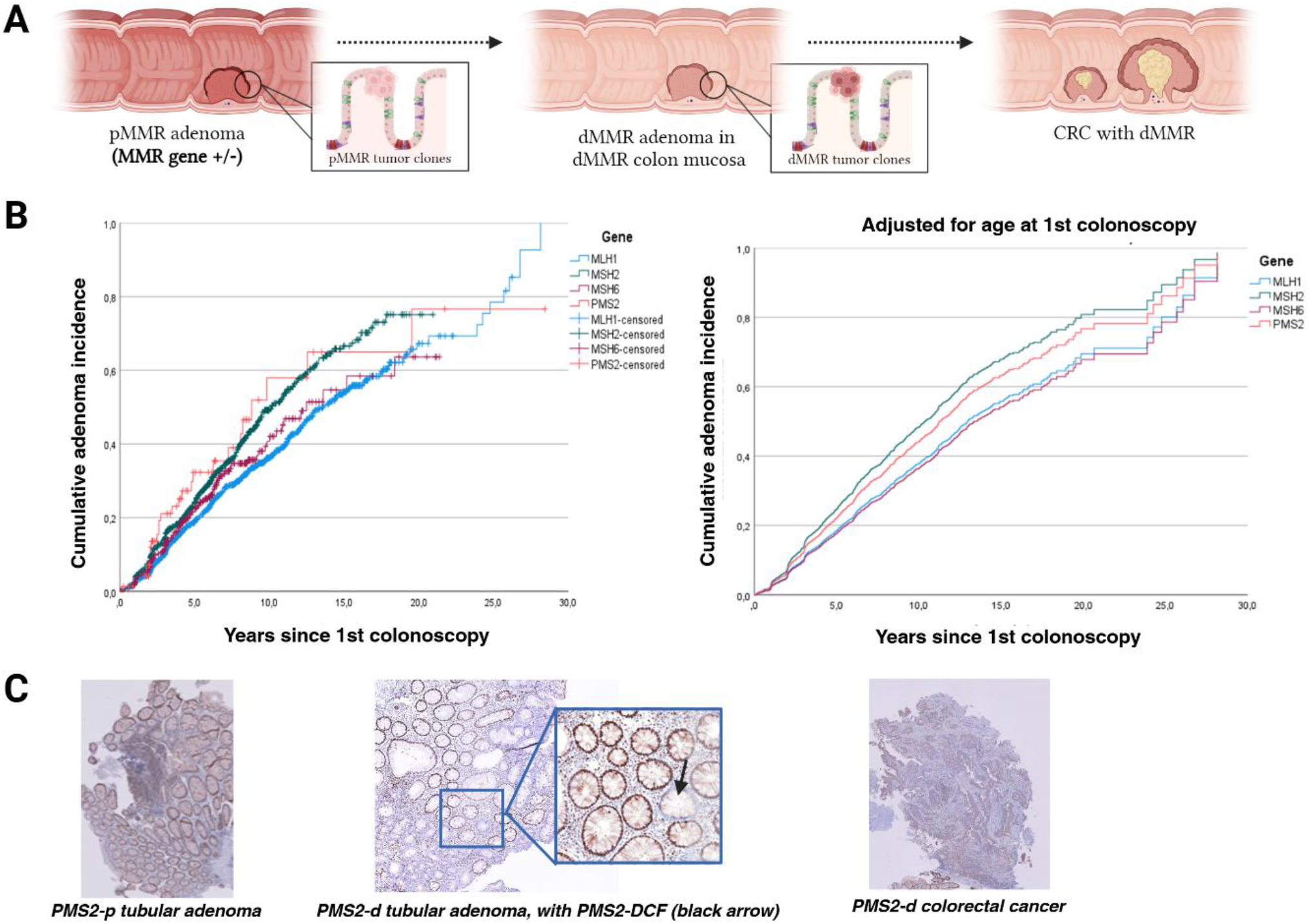
**A)** PMS2 Lynch syndrome carcinogenesis pathway, in which MMR capacity is lost over time as the adenoma progresses into colorectal cancer, **B)** Cumulative incidence of adenomas among four Lynch syndrome-associated genes (left) and after adjusting for age at first colonoscopy (right), and **C)** Immunohistochemical staining patterns of adenomatous lesions: PMS2-proficient tubular adenoma, PMS2-deficient tubular adenoma with PMS2-DCF, PMS2-deficient colorectal cancer

The Dutch *PMS2* cohort consists of 171 individuals, its clinical characteristics summarized in Table 1. A total of 576 colonoscopies were performed between 1987 and 2023, representing 1769 years of follow-up. The cumulative incidence of *PMS2*-associated adenomas 10 years after index colonoscopy was 58% (39.2%-81.2%), highest among the four MMR genes followed by *MSH2*, as demonstrated in Figure 1B (left panel). A previous report by Engel et al.^7^ demonstrated adenoma incidence rates of 44.2% for *MSH2*, 38.4% for *MSH6*, and 32.2% for *MLH1*. We were able to merge this data with our *PMS2* cohort, allowing us to compare the different Lynch syndrome subtypes. Because of differences in mean age at index colonoscopy (Table 1) which could impact the time until the occurrence of an adenoma, we performed a Cox regression analysis. Age at first surveillance was indeed associated with the risk of developing adenomas in all MMR gene mutation carriers (HR = 1.027, 95% CI: 1.022-1.32, p <0.001). Additionally, we added gender to the analysis and found that female mutation carriers have a lower hazard of adenoma development compared to males (HR = 0.781, 95% CI: 0.689-0.879 p <0.001). The analyses showed similar directional trends for the cumulative incidence across gene groups in both Kaplan-Meier analysis and the adjusted Cox regression model, with the highest risk for *MSH2* PV carriers, followed by *PMS2-* and the lowest risk for *MSH6*- and *MLH1*- PV carriers (Figure 1B, right panel).

**Table 1.** Clinical and histopathological characteristics of Dutch *PMS2* cohort.

| **Clinical characteristics** |  |  |
| --- | --- | --- |
| Sex | Female: 102 (59.6%)<br>Male: 69 (40.4%) |  |
| Mean age at first/index colonoscopy (range) | *MLH1*: 42.6 years (18.2-85.8 years)<br>*MSH2*: 44.1 years (18.9-79.1 years)<br>*MSH6*: 48.8 years (20.4-83 years)<br>*PMS2*: 50.6 years (19.9-85.6 years) |  |
| Mean follow-up time (range) | *MLH1*: 10.2 years (0.5-31.1 years)<br>*MSH2*: 8.8 years (0.6-22.7 years)<br>*MSH6*: 7.4 years (0.6-21.4 years)<br>*PMS2*: 6.1 years (0-28.4 years) |  |
| Adenoma incidence rates*# | *MLH1*: 36.3%, 95% CI = 33.2-39.4%<br>*MSH2*: 49.5%, 95% CI = 41.6-53.8%<br>*MSH6*: 40.7%, 95% CI = 33.1-48.3%<br>*PMS2*: 58%, 95% CI = 39.2-81.2% |  |
| **Histopathological features<sup>§</sup>** | PMS2-proficient | PMS2-deficient |
| Hyperplastic polyp | 8 (6.5%) | 0 |
| Sessile serrated adenoma | 1 (0.08%) | 0 |
| Tubular adenoma | 94 (76.4%) | 15 (12.2%) |
| Tubulovillous adenoma | 2 (0.16%) | 1 (0.08%) |
| Colorectal cancer | 0 | 2 (0.16%) |

\*Data differs slightly with that reported by Engel et al, due to exclusion of one center.<sup>7</sup>;

Next, we set out to investigate the prevalence of PMS2 deficiency in (early) lesions detected in the PMS2-cohort and conducted immunohistochemical (IHC) staining for anti-MLH1 and anti-PMS2 antibodies on 123 paraffin blocks containing lesions removed during colonoscopy between 2018-2023. See supplementary data for a detailed description of the methods. The morphological features of the lesions are described in Table 1. Of the 123 IHC-stained lesions 109 were tubular adenomas, 86.2% of which (n=94/109) retained PMS2 protein expression. As expected for germline *PMS2* PV carriers, MLH1 staining remained intact in all adenomas. The finding that only 13.8% of adenomas removed in our PMS2 cohort were MMR deficient is in striking contrast with a meta-analysis involving 640 adenomas from *MLH1, MSH2*, and *MSH6* PV carriers of which 76.7% of dysplastic adenomas were already MMR-deficient (n = 491, 95% CI: 73.3–79.8%)^5^.

Our observations support the notion that loss of the *PMS2* wildtype allele, and thus occurrence of PMS2-defiency, may occur later in the adenoma-carcinoma evolution in *PMS2* PV carriers. Our analyses also demonstrated that despite rare occurrence of *PMS2*-CRC among our cohort (n=2), 58% of *PMS2* variant carriers developed adenomas within 10 years of first colonoscopy. This means that most adenomas in patients with PMS2-LS likely do not (rapidly) progress to CRC. Furthermore, the high cumulative PMS2 adenoma incidence, coupled with low PMS2-CRC occurrence, suggests that routine surveillance colonoscopy was successful to prevent almost all occurrences of CRC. Our current data combining epidemiological and IHC analyses corroborate the preference of an “accelerated” adenoma-carcinoma sequence in PMS2-Lynch syndrome: early PMS2 adenomas are MMR-proficient and a minority progressively lose MMR capacity over time which sets the acceleration in motion.

An intriguing observation is that the incidence of adenomas in PMS2 PV carriers appears similar to that of *MSH2* PV carriers, despite the hypothesis that the latter subtype develops CRC from MMRd crypt foci through an intermittent adenoma phase. This alternative route may contribute to the relatively high adenoma incidence seen in MSH2-versus MLH1-associated Lynch syndrome patients, as MSH2-deficency drives the development of an adenoma. A possible explanation for the comparable adenoma incidence observed in *PMS2* PV carriers could be the presence of additional, non-MMR-related risk factors in our clinically-ascertained cohort, indirectly increasing their risk of CRC. Beside the underlying germline *PMS2* PV, such factors that innately confer higher risk (both external and internal) of adenoma development remain to be investigated. Additionally, the older age of our cohort of PMS2 PV carriers may also increase the risk of adenoma detection due to mutations that accumulate in the colonic epithelium over time, although the age-related mutation accumulation is not enough to induce the loss of *PMS2* heterozygosity as demonstrated by the overwhelming majority of PMS2-proficient adenomas among our cohort.

To the best of our knowledge, our study is the first to thoroughly assess adenoma morphology and PMS2 deficiency status based on immunohistochemistry. Our data can be combined with similar PMS2-focused data from other countries, as well as sequencing data from both normal-looking crypts and adenomas of *PMS2* PV carriers, in order to paint a complete picture of its early-stage carcinogenesis. Future studies should investigate the varying “mutational evolution” of events leading to MMR deficiency, adenoma development, and/or progression into LS-CRC. By understanding the early molecular alterations for each LS-associated gene, prevention approaches (e.g. neoantigen-based vaccinations^9^ or NSAID-based chemoprevention^10^) can be tailored to each molecular pathway.

In conclusion, PMS2-deficiency is rare in (early) adenomas of *PMS2* PV carriers, despite frequent adenoma detection after index colonoscopy. Adenoma formation in this Lynch syndrome subtype thus appears largely independent of PMS2 loss, highlighting the need to uncover other key drivers to guide effective prevention.

## Data Availability

All data produced in the present study are available in the manuscript, otherwise will be made available upon reasonable request to the authors.

## Funding

The LYNCH-GPS project is funded by the Dutch Cancer Society Young Investigator Grant (KWF 2019-12570). We also thank (patients who voluntarily participate in LYNCH-GPS study, PALGA, MSc. Biomedical Sciences intern A.M. Reilly, pathology archive manager H.J. Hilbrands, pathology technicians, external collaborators from various hospitals in the Netherlands) for providing the FFPE material and assisting us in sample storage and/or processing.

## Ethical statement

This study was carried out following the ethical approval issued by the Institutional Review Board of Leiden University Medical Centre (No. P01.019) in a previous project.

## Conflict of interest

The authors declare no competing conflicts of interest.

## Notes

### Competing Interest Statement

The authors have declared no competing interest.

