## Supplementary materials for "Adenomas Are Frequent in PMS2 Lynch Syndrome patients but Rarely Mismatch Repair Deficient"

1. **Materials and Methods:**

a. *Data collection and ethical statement*

This study was carried out following the ethical approval issued by the Institutional Review Board of Leiden University Medical Centre (No. P01.019) in a previous project (UL 2012-5515) and was approved by the Medical Ethics Committee of the University Medical Centre Groningen. Consent was obtained to request clinical information and pathology samples for 186 Dutch LS patients with a confirmed pathogenic germline *PMS2* variant diagnosed at Dutch family cancer clinics. Obtaining pathology reports was facilitated by PALGA, the nationwide network and registry of histology and cytopathology in the Netherlands. As PALGA encompasses all pathology laboratories in the Netherlands, all pathology reports on each patient can be obtained, even if a patient attended different hospitals for colonoscopies. Corresponding colonoscopy reports were requested at the respective gastroenterology departments. For 15 *PMS2* variant carriers both the PALGA search and request for colonoscopy reports did not yield any results, it is most likely that these patients are not undergoing regular surveillance. Therefore, they were excluded from our study and we requested available materials from 171 pathogenic *PMS2* variant carriers. Specimen collection and experiments were conducted according to the medical ethical guidelines described in the Declaration of Helsinki.

b. *PMS2 variant analysis*

Our cohort consisted of clinically ascertained families in which variant analysis was initiated due to (histological) pre-screening by immunohistochemistry and/or microsatellite instability, usually because a family met the Bethesda criteria (Umar et al. 2004). Germline *PMS2* variant screening was performed as previously described (ten Broeke et al. 2015; Ten Broeke, van der Klift, et al. 2018; van der Klift et al. 2016). Comprehensive strategies were applied to avoid unreliable variant detection caused by interference from pseudogene sequences and frequent gene conversion events (van der Klift et al. 2016). All variants found in our cohort are listed in Supplemental Table 1.

c. *Immunohistochemistry for MLH1-PMS2 heterodimer*

We retrieved 123 formalin-fixed, paraffin-embedded (FFPE) tissue blocks from *PMS2* variant carriers that contained adenomas and were isolated between 2018 and 2023. We performed immunohistochemical (IHC) analysis of PMS2 and MLH1 protein expression. In brief, the FFPE materials were sectioned at 4-5 μm thickness and mounted on adhesive microscope slides (Epredia SuperFrost Plus Adhesion slides, Fisher Scientific, United Kingdom). The glass slides were loaded to Benchmark ULTRA automated stainer (Roche Diagnostics, USA) where samples were stained with anti-PMS2 antibody (Ventana anti-PMS2 A16-4 mouse monoclonal primary antibody, Roche Diagnostics, USA) and anti-MLH1 antibody (Ventana anti-MLH1 M1 mouse monoclonal primary antibody, Roche Diagnostics, USA) according to manufacturer's instructions. Secondary antibody was used to aid visualisation of PMS2/MLH1 heterodimer (Optiview DAB IHC Detection Kit, Roche Diagnostics, USA). Following incubation of primary and secondary antibodies, the slides were washed, dehydrated, and counterstained by hematoxylin.

Dark brown nuclear staining indicates the presence of either MLH1 or PMS2 in stromal cells and lymphocytes, this staining pattern is then considered as internal control. Colonic epithelium without nuclear staining of PMS2 or MLH1 but intact staining of internal control is deemed as PMS2-deficient or MLH1-deficient, respectively. Adenomas and advanced adenomas were classified according to the Vienna classification of gastrointestinal epithelial neoplasia (Schlemper et al. 2000) and more recent WHO classification of digestive system tumours (*WHO Classification of Tumours. Digestive*

*System Tumours: WHO Classification of Tumours, Volume 1* 2019). Advanced adenomas were defined by a size of ≥1 cm in diameter, a villous component of >25%, and/or the presence of high-grade dysplasia. Interpretation of IHC results were done by PhD student (K.D.A) under supervision of an experienced pathologist (G.K-U).

d. *Statistical analysis*

Descriptive results of colonoscopy findings were computed using SPSS (version 28). A Kaplan Meier analysis was carried out to estimate time to first adenoma or first advanced adenoma. Cox regression analyses were conducted to assess the influence of biological sex and the age of index colonoscopy on adenoma incidence. Both K.D.A and L.L conducted the Kaplan-Meier and Cox regression analyses. Syntax for both analyses can be found in Appendix 1. Results were compared to data from a study by Engel et al., in which cumulative adenoma incidence rates of *MLH1*-, *MSH2*-, and *MSH6* pathogenic variant carriers were reported (Engel et al., 2020).

**Supplementary Table 1: List of pathogenic variants encountered in the Dutch PMS2 cohort**

| Series | Sample number | Origin | Sex | Genotype (m) | Genotype (a) |
|---|---|---|---|---|---|
| R23-800681 | I-1 | HAGA | M | c.150delinsAG; exon 2 | N/A |
| | II-1 | | M | c.150delinsAG; exon 2 | N/A |
| | III-1 | | M | c.1882C>T | p.Arg628* |
| | IV-1 | | M | c.1882C>T | p.Arg628* |
| | V-1 | | M | c.150delinsAG; exon 2 | N/A |
| | VI-1 | | M | c.150delinsAG; exon 2 | N/A |
| | VII-1 | | M | c.150delinsAG; exon 2 | N/A |
| | VIII-1 | | M | c.1882C>T | p.Arg628* |
| | IX-1 | | M | c.1882C>T | p.Arg628* |
| | XI-1 | | M | c.1882C>T | p.Arg628* |
| | X-1 | | M | c.1882C>T | p.Arg628* |
| R23-800757 | I-1 | AMC | F | deletion exon 11-15 | N/A |
| | II-1 | | F | deletion exon 11-15 | N/A |
| | III-1 | | M | c.1831_1832insA | N/A |
| | IV-1 | | M | c.1831_1832insA | N/A |
| | V-1 | | M | c.1831_1832insA | N/A |
| | VI-1 | | M | c.1831_1832insA | N/A |
| | VII-1 | | M | c.1831_1832insA | N/A |
| | VIII-1 | | F | deletion exon 14 | N/A |
| | IX-1 | | F | deletion exon 11-15 | N/A |
| R23-800836 | I-1 | Rijnstate | M | c.2192_2196del | p.Leu731Cysfs*3 |
| | II-1 | | M | c.2192_2196del | p.Leu731Cysfs*3 |
| | III-1 | | M | c.2192_2196del | p.Leu731Cysfs*3 |
| | IV-1 | | M | c.2192_2196del | p.Leu731Cysfs*3 |
| | V-1 | | M | c.2192_2196del | p.Leu731Cysfs*3 |
| | VI-1 | | M | c.2192_2196del | p.Leu731Cysfs*3 |
| | VII-1 | | M | c.2192_2196del | p.Leu731Cysfs*3 |
| R23-800837 | I-1 | Symbiant | F | deletie exon 11-12 | N/A |
| | I-2 | | F | deletie exon 11-12 | N/A |

| | | | | | |
|---|---|---|---|---|---|
| | II-1 | | F | c.697C>T | p.Gln233* |
| | III-1 | | F | c.697C>T | p.Gln233* |
| | IV-1 | | F | deletie exon 11-12 | N/A |
| | V-1 | | F | deletie exon 11-12 | N/A |
| | VI-1 | | F | deletie exon 11-12 | N/A |
| R23-800873 | | St. Antonius | M | c.1882C>T | p.Arg628X |
| R23-800963 | I-1 | Maastricht | F | c.24-12_107delinsAAAT | intron mutatie |
| | II-1 | | F | c.24-12_107delinsAAAT | intron mutatie |
| R23-800964 | I-1 | Martini | F | c.736_741delCCCCCTinsTGTGTGTGAAG | p.Pro246CysfsX3 |
| | II-1 | | M | c.325dup | p.Glu109Glyfs*30 |
| | IV-1 | | F | c. 856_857del | p.Asp286Glnfs*12 |
| | V-1 | | F | c. 856_857del | p.Asp286Glnfs*12 |
| R23-801111 | I-1 | Radboud | F | c.2404C>T | p.Arg802X |
| | II-1 | | M | deletion exon 10 (c.989-?_1144+?del) | |
| | III-1 | | F | c.354-1G>A | intron |
| | IV-1 | | M | c.1882C>T | p.Arg628* |
| | V-1 | | M | c.1882C>T | p.Arg628* |
| | VI-1 | | M | c.1882C>T | N/A |
| | VII-1 | | F | c.2404C>T | p.Arg802X |
| | VIII-1 | | F | c.1882C>T | p.Arg628* |
| | IX-1 | | M | c.1882C>T | p.Arg628* |
| | X-1 | | M | deletion exon 10 (c.989-?_1144+?del) | N/A |
| | XI-1 | | M | c.1882C>T | p.Arg628* |
| | XII-1 | | F | c.1882C>T | p.Arg628* |
| R24-800094 | I-1 | OLVG | M | N/A | N/A |
| | II-1 | | F | c.804-60_804-59insJN866832.1 | N/A |
| | III-1 | | M | N/A | N/A |
| | IV-1 | | M | N/A | N/A |
| | V-1 | | F | c.804-60_804-59insJN866832.1 | N/A |
| | VI-1 | | M | N/A | N/A |
| | VII-1 | | M | N/A | N/A |
| R24-800153 | I-1 | Pathologie Friesland | M | deletion exon 3-7 | N/A |
| | II-1 | | M | deletion exon 3-7 | N/A |
| | III-1 | | F | deletion exon 3-7 | N/A |
| | IV-1 | | F | c.736_741delinsTGTGTGTGAAG | p.Pro246Cysfs*3 |
| | V-1 | | F | c.736_741delCCCCCTinsTGTGTGTGAAG | p.Pro246CysfsX3 |
| | VI-1 | | F | c.736_741delCCCCCTinsTGTGTGTGAAG | p.Pro246CysfsX3 |
| | VII-1 | | F | c.736_741delCCCCCTinsTGTGTGTGAAG | p.Pro246CysfsX3 |
| | VIII-1 | | F | c.736_741delCCCCCTinsTGTGTGTGAAG | p.Pro246CysfsX3 |
| | IX-1 | | F | c.736_741delinsTGTGTGTGAAG | p.Pro246Cysfs*3 |
| R24-800212 | I-1 | LUMC | F | c.325dupG | p.Glu109fs* |
| | II-1 | | F | c.1882C>T | p.Arg628* |

| | | | | | |
|---|---|---|---|---|---|
| | III-2 | | F | deletion exon 11-15 | N/A |
| | IV-1 | | F | c.736_741delCCCCCTinsTGTGTGTGAAG | p.Pro246CysfsX3 |
| | V-1 | | F | deletion exon 11-15 | N/A |
| | V-2 | | F | deletion exon 11-15 | N/A |
| | VI-1 | | F | c.325dupG | p.Glu109fs* |
| | VII-1 | | F | c.325dupG | p.Glu109fs* |
| | VIII-1 | | F | deletion exon 11-15 | N/A |
| | IX-1 | | F | c.736_741delCCCCCTinsTGTGTGTGAAG | p.Pro246CysfsX3 |
| R24-800436 | I-1 | Haaglanden | M | deletion exon 5 t/m 7 | N/A |
| | II-1 | | M | deletion exon 5 t/m 7 | N/A |
| | III-1 | | M | deletion exon 5 t/m 7 | N/A |

**Appendix 1: Syntaxes for Kaplan-Meier and Cox regression analyses**

1. **Case selection:**

IF (MISSING(years_since_1st_scopy) AND adenomas = 0)

years_since_1st_scopy = (age_last_scopy - age_1st_scopy).

EXECUTE.

IF (adenomas=1 AND Gene=4) years_since_1st_scopy=(age_1st_scopy_adenoma-age_1st_scopy).

EXECUTE.

IF (crcage < age_last_scopy) years_since_1st_scopy=(crcage-age_1st_scopy).

EXECUTE.

IF (age_1st_scopy_adenoma>crcage) adenomas=0.

EXECUTE.

2. **Life table for all cases**

TEMPORARY.

SELECT IF filter_crc=1 and filter_scopy=1 and filter_adenoma=1.

SURVIVAL TABLE=years_since_1st_scopy

/INTERVAL=THRU 20 BY 5

/STATUS=adenomas(1)

/PRINT=TABLE

/PLOTS (OMS)=years_since_1st_scopy.

**Life Table[a]**

| Interval Start Time | Number Entering Interval | Number Withdrawing during Interval | Number Exposed to Risk | Number of Terminal Events | Proportion Terminating | Proportion Surviving | Cumulative Proportion Surviving at End of Interval | Std. Error of Cumulative Proportion Surviving at End of Interval | Probability Density | Std. Error of Probability Density | Hazard Rate | Std. Error of Hazard Rate |
|---|---|---|---|---|---|---|---|---|---|---|---|---|
| 0 | 2644 | 578 | 2355,000 | 500 | ,21 | ,79 | ,79 | ,01 | ,042 | ,002 | ,05 | ,00 |
| 5 | 1566 | 628 | 1252,000 | 314 | ,25 | ,75 | ,59 | ,01 | ,040 | ,002 | ,06 | ,00 |
| 10 | 624 | 285 | 481,500 | 135 | ,28 | ,72 | ,42 | ,01 | ,033 | ,003 | ,07 | ,01 |
| 15 | 204 | 146 | 131,000 | 29 | ,22 | ,78 | ,33 | ,02 | ,019 | ,003 | ,05 | ,01 |

a. The median survival time is 12,72380109464231

### 3. Kaplan meier analysis

```
TEMPORARY.

SELECT IF filter_crc=1 AND filter_scopy=1 AND filter_adenoma=1.

KM years_since_first_scopy BY gene

 /STATUS=adenomas(1)

 /PRINT TABLE MEANS

 /PLOT OMS

 /TEST LOGRANK

/COMPARE OVERALL POOLED.
```

**Case Processing Summary**

| Gene | Total N | N of Events | Censored | |
|---|---|---|---|---|
| | | | N | Percent |
| MLH1 | 1324 | 489 | 835 | 63,1% |
| MSH2 | 911 | 371 | 540 | 59,3% |
| MSH6 | 334 | 100 | 234 | 70,1% |
| PMS2 | 75 | 26 | 49 | 65,3% |
| Overall | 2644 | 986 | 1658 | 62,7% |

**Means and Medians for Survival Time**

| Gene | Mean[a] | | | Median | | |
|---|---|---|---|---|---|---|
| | Estimate | Std. Error | 95% Confidence Interval | Estimate | Std. Error | 95% Confidence Interval |

| | | | Lower Bound | Upper Bound | | | Lower Bound | Upper Bound |
|---|---|---|---|---|---|---|---|---|
| MLH1 | 14,761 | ,419 | 13,938 | 15,583 | 13,600 | ,587 | 12,449 | 14,751 |
| MSH2 | 11,246 | ,329 | 10,601 | 11,892 | 10,140 | ,495 | 9,170 | 11,110 |
| MSH6 | 12,922 | ,687 | 11,576 | 14,268 | 12,480 | 1,643 | 9,260 | 15,700 |
| PMS2 | 12,834 | 1,996 | 8,921 | 16,746 | 8,816 | 1,040 | 6.777 | 10,854 |
| Overall | 13,976 | ,331 | 13,327 | 14,626 | 12,200 | ,350 | 11,513 | 12,887 |

a. Estimation is limited to the largest survival time if it is censored.

**Overall Comparisons**

| | Chi-Square | Df | Sig. |
|---|---|---|---|
| Log Rank (Mantel-Cox) | 24,693 | 3 | <,001 |

Test of equality of survival distributions for the different levels of Gene.

| Gene | Year | Survival est. | SE | Cumulative Incidence | Upper CI cum | Lower CI cum |
|---|---|---|---|---|---|---|
| **MLH1** | 9.9 | 0.637 | 0.016 | 0.363 (36.3%) | 0.394 (39.4%) | 0.332 (33.2%) |
| **MSH2** | 10.0 | 0.505 | 0.022 | 0.495 (49.5%) | 0.538 (53.8%) | 0.416 (41.6%) |
| **MSH6** | 9.85 | 0.593 | 0.039 | 0.407 (40.7%) | 0.483 (48.3%) | 0.331 (33.1%) |
| **PMS2** | 9.83 | 0.420 | 0.096 | 0.580 (58.0%) | 0.812 (81.2%) | 0.392 (39.2%) |

***Comparison to Engel et al. data:***

| Source | MLH1 (95% CI) | MSH2 (95% CI) | MSH6 (95% CI) | PMS2 (95% CI) |
|---|---|---|---|---|
| Engel et al. (2020) | 32.2% (29.2-35.2%) | 44.2% (40-48.4%) | 38.4% (30.8-45.9%) | N/A |
| PMS2 data added | 36.3% (33.2%-39.4%) | 49.5% (41.6%-53.8%) | 40.7% (33.1%-48.3%) | 58.0% (39.2%-81.2%) |

4. **<u>Cox regression analysis, covariates = gene, sex, age</u>**

```
TEMPORARY.
SELECT IF filter_crc=1 and filter_scopy=1 and filter_adenoma=1.
COXREG years_since_1st_scopy
  /STATUS=adenomas(1)
  /PATTERN BY Gene
  /CONTRAST (sex)=Indicator(1)
  /CONTRAST (Gene)=Indicator(4)
  /METHOD=ENTER Gene age_1st_scopy sex
  /PLOT OMS
```

```
/PRINT=CI(95)
/CRITERIA=PIN(.05) POUT(.10) ITERATE(20).
```

**Case Processing Summary**

| | | N | Percent |
|---|---|---|---|
| Cases available in analysis | Event[a] | 986 | 37,3% |
| | Censored | 1629 | 61,6% |
| | Total | 2615 | 98,9% |
| Cases dropped | Cases with missing values | 0 | 0,0% |
| | Cases with negative time | 0 | 0,0% |
| | Censored cases before the earliest event in a stratum | 29 | 1,1% |
| | Total | 29 | 1,1% |
| Total | | 2644 | 100,0% |

a. Dependent Variable: years_since_1st_scopy

**Categorical Variable Codings[a,c]**

| | | Frequency | (1) | (2) | (3) |
|---|---|---|---|---|---|
| sex[b] | 1 | 1252 | 0 | | |
| | 2 | 1392 | 1 | | |
| Gene[b] | 1=MLH1 | 1324 | 1 | 0 | 0 |
| | 2=MSH2 | 911 | 0 | 1 | 0 |
| | 3=MSH6 | 334 | 0 | 0 | 1 |
| | 4=PMS2 | 75 | 0 | 0 | 0 |

a. Category variable: sex

b. Indicator Parameter Coding

c. Category variable: Gene

**Variables in the Equation**

| | | | | | | | 95,0% CI for Exp(B) | |
|---|---|---|---|---|---|---|---|---|
| | B | SE | Wald | df | Sig. | Exp(B) | Lower | Upper |
| Gene | | | 25,903 | 3 | <,001 | | | |
| Gene(1) | -,204 | ,205 | ,993 | 1 | ,319 | ,816 | ,546 | 1,218 |
| Gene(2) | ,126 | ,207 | ,371 | 1 | ,542 | 1,134 | ,756 | 1,701 |

| | | | | | | | | |
|---|---|---|---|---|---|---|---|---|
| Gene(3) | -,250 | ,223 | 1,257 | 1 | ,262 | ,779 | ,503 | 1,206 |
| age_1st_scopy | ,027 | ,003 | 116,079 | 1 | <,001 | 1,027 | 1,022 | 1,032 |
| sex | -,247 | ,064 | 14,927 | 1 | <,001 | ,781 | ,690 | ,886 |
